# Feasibility of Mini Group Interpersonal psychotherapy (IPT-G mini) for adolescents impacted by COVID-19 in East Africa

**DOI:** 10.1101/2025.06.13.25329611

**Authors:** Obadia Yator, Shillah M. Mwavua, Albert Tele, Joseph Kathono, Vincent Nyongesa, Nabila Amin, Manasi Kumar

**Author notes:** **Corresponding author:** (MK).

## Abstract

**Introduction:** Group Interpersonal Psychotherapy (IPT-G) has universal elements that appeal to non-Western cultural contexts, alongside offering flexibility to make contextual adaptations and making it an intervention of choice for adolescent populations in LMICs. We anticipate that a shorter four-session version, which we are calling IPT-G Mini, offered to adolescents and young people during the COVID-19 pandemic would effectively mitigate psychological distress.

**Materials and methods:** IPT-G mini was offered by lay health workers to 40 adolescents aged 10-19 years in Nairobi, Kenya, experiencing depressive, stress, and anxiety symptoms during the COVID-19 epidemic in June-September 2021. Primary outcomes were a reduction in depression, and psychological distress on the Patient Health Questionnaire (PHQ-9), Revised Children’s Anxiety, and Kessler psychological distress (K10) scales.

**Results:** The intervention was effective, with a significant reduction in psychological distress (K-10) and depression (PHQ-9) scores at the end of the intervention (with p<0.05) Cohen’s d=0.49 (95% C.I. =0.04-0.94) and d=0.51 (95% C.I. = 0.06-0.96) respectively. Younger participants (10-14 years) had better outcomes than older ones on K10 and PHQ-9 assessments.

**Conclusion:** IPT-G mini must be tested in a more controlled setting with a larger and more diverse sample. It does appear to reduce distress and depressive symptoms, and further evaluation may be needed to refine the adapted version. Despite the challenges brought by the pandemic, the timely and effective intervention focused on interpersonal issues proved beneficial to participants. Addressing various problem areas helped provide perspective and support in dealing with multiple losses and socioeconomic hardships experienced by our young participants.

**Trial registry:** Pan African Clinical Trial Registry (pactr.samrc.ac.za) PACTR202501888041900

## Introduction

Mental health disorders are highly prevalent in lower-and-middle-income countries (LMIC) [1] and tailored mental health services are not only underdeveloped but underfunded within health system as well. The 2019 GBD data for adolescents ages 10-24 years in Kenya observed the highest prevalence of mental disorders in Depressive, Anxiety, and Conduct Disorders [2]. Studies have also underscored the discrepancy between the demands and availability of mental healthcare for adolescents and youth populations throughout the world [3,4]. Additionally, the COVID-19 pandemic further worsened the detrimental effects on children and adolescents due to lockdown, disrupted schooling, adversely impacted learning outcomes, and decreased opportunities for socialization and recreation [5].

The World Health Organization (WHO) has recognized Interpersonal Psychotherapy (IPT) as a key evidence-based intervention [6,7] for task-shared and task-shifted mental health delivery in low-resource contexts. In this sense, it is the redistribution and delegation of activities and tasks among health workforce teams, where both fully trained and qualified health workers, alongside other non-specialist or lay health workers with shorter training duration, work together to deliver mental health services [8].The WHO’s Mental Health Gap Action Program (mhGAP) [9] and its associated quality and service improvement guidelines promote the idea of quick assessment, management, and referral pipeline in primary and tertiary care. IPT is one of the recommended treatments within WHO mhGAP.

Several studies have used IPT during the pandemic [10–12]. A case report by Hu et al, documented that 4 sessions of IPT delivered face-to-face to a patient diagnosed with depression, anxiety, and COVID-19, led to a reduction in his depression symptoms and anxiety evidenced by a decrease in assessment scores using the Hamilton rating scale and PHQ-9 to measure depression and anxiety levels, after the therapy [10]. Another study by Swartz et al. in 2014 found that the concentrated focus on Brief Interpersonal Psychotherapy (IPT-B) is highly beneficial for individuals dealing with various interpersonal issues. The framework of IPT-B compels patients (and therapists) to target the most urgent—and ideally resolvable— interpersonal problem as the core of the treatment. This approach offers a sense of direction and structure amidst the turmoil [13]. Considering Kenyan adolescents, Kumar et al., developed a model for implementing IPT within the WHO’s Mental Health Gap Action Program (known as mhGAP)-a depression care framework for pregnant adolescents. The authors enhanced the existing 8-session version in their model by incorporating additional implementation strategies like stigma reduction, task-sharing, and peer engagement. Additionally, they tailored a shorter 4-session version called IPT-G Mini to effectively address perinatal depression among this population [14].

The protocol for this mini IPT was published a few years back [15,16], and the publications highlighted the possible benefits of task shifting in addressing mental health problems within low-resource settings in Kenya and demonstrated that delivery of IPT-G was acceptable and feasible by lay and non-specialist health workers to adolescents receiving primary care services [15]. Mattos et al., concluded that implementing IPT-G in primary care is a complex process involving several steps. In the first step, successful training of health professionals is crucial; the subsequent steps are more complex and, therefore, require careful planning to maximize the success of an integrated treatment within primary care settings [13,16–18].

### Addressing the needs of varied adolescent subgroups in community-based interventions during the pandemic

This study was embedded under a UNICEF Measurement of Adolescent Mental Health at Population Level [19], where we worked with Nairobi County and the Ministry of Health’s Division of Mental Health and Substance Use, to test population-level measurement of key mental health tools like PHQ-9, [20] Revised Children’s Anxiety Depression Scale (RCADS), [21]. UNICEF Multiple Indicator Cluster Survey (MICS) [22] assessment for adolescent functioning [23,24]. The participants who were screened with high depression and anxiety or found to have suicidal thoughts, intent, and actions were provided referrals to the County and National referral hospitals available in Nairobi. We also included high-risk participants, such as those who lost family members due to COVID-19, as they were expressing distress and need for help. The pandemic caused a lot of disruption in routine family activities, affecting the flow of income, food security, boredom due to lockdown, unusual conflicts following unmet needs within the families, anticipated grief in some instances arising from panic from the disturbing scenes on social media, and an increase in teenage pregnancy, hence in this context, we decided to deploy IPT-G mini which is a 4 sessions therapy model.

## Methods

### Participants, setting, and tools

A survey on the mental health needs of adolescents ages 10-19 years was carried out from June to July 2021 in the two primary care clinic sites (n=250) where large informal settlement neighborhoods are based. This survey was a UNICEF’s Measurement of Mental Health among Adolescents at the Population Level (MMAP) initiative [25] that was embedded under a larger study on mental health interventions for pregnant adolescents (INSPIRE) study [15]. Participants who had a score of moderate to severe depression (PHQ-9 score of 15 to 27) or anxiety or had suicidal ideations were invited to participate in the intervention (n=40). Participants above 18 years gave consent, while those below 18 years gave assent, followed by their parent or caregiver consent. The intervention was delivered in the primary health care clinic in Nairobi, which was accessible and closer to the participants’ homes.

We used the mental health needs of adolescents’ survey as baseline screening information to recruit participants for this rapid trial and follow-up that happened between August and October 2021. The baseline survey collected socio-demographic information, depression, anxiety, and social functioning. This assessment was conducted by lay health workers (also known as community health promoters). These are not formally trained as health care professionals but belong to and possess both cultural and linguistic understanding of the community they serve, thus playing a crucial role in promoting and improving community health. The lay health workers were trained on identifying participants, assenting and consenting, conducting research with human subjects, and administering the measures used in the baseline survey item by item. A few participants were invited, and community health promoters piloted the tools under the close supervision of the research team. Post-intervention (after delivery of 4 sessions) data was collected using a recently validated PHQ-9 scale by Tele et al., which established that the psychometric properties of both English and Swahili-adapted versions were comparable indications they were reliable and valid instruments for detecting major depressive disorders among adolescents [22]. Depression is a clinical diagnosis characterized by a prolonged, persistent state of low mood, while psychological distress, as assessed using K10 [26], is a broader domain encompassing a range of emotional and mental health symptoms that may not meet the criteria for a specific mental health disorder (Fig 1).

**Fig 1.**
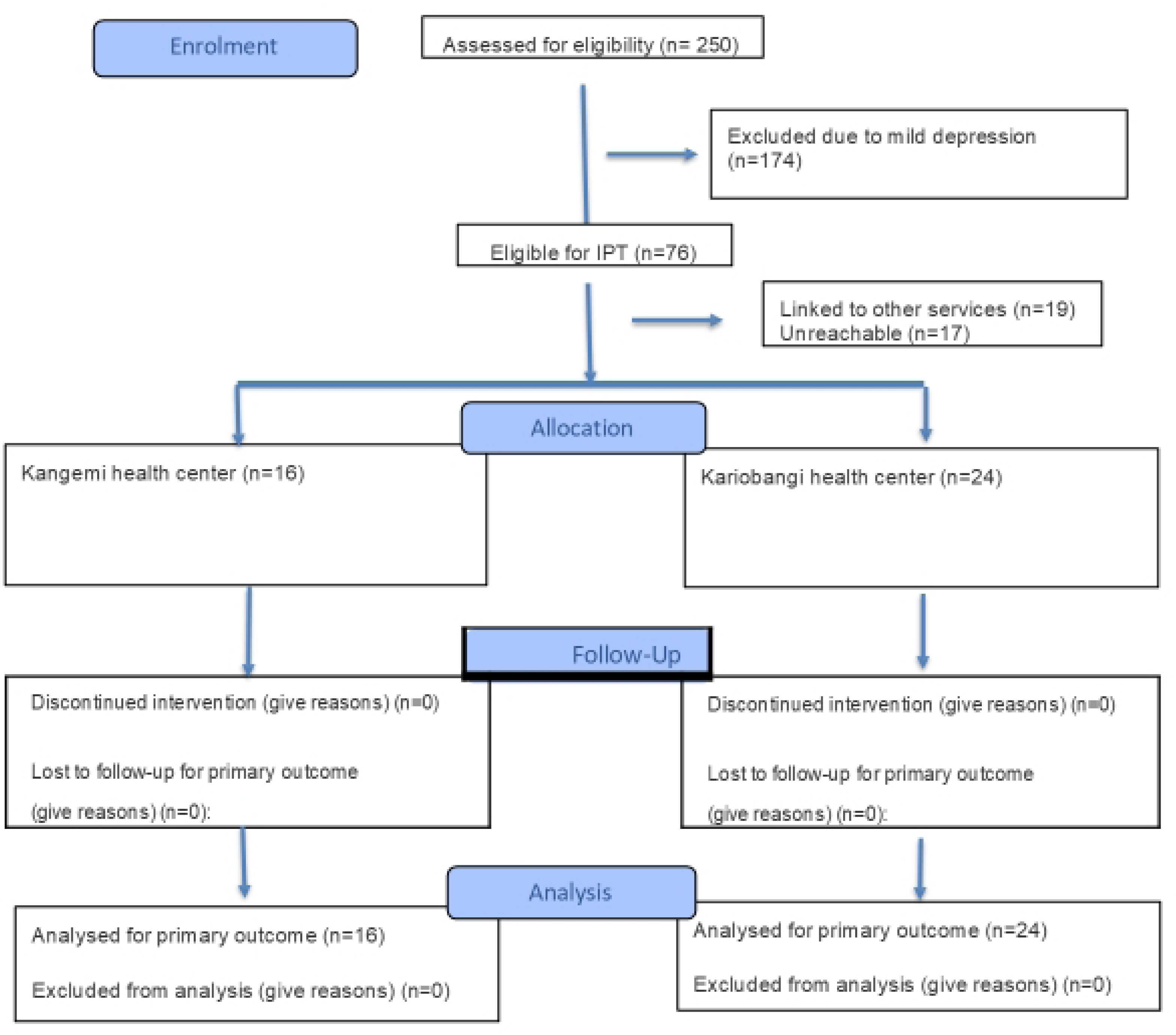
CONSORT flowchart.

### Kariobangi and Kangemi Health Centers

Kariobangi health center is a level three facility managed by Nairobi City County Government. Level three health facilities include health centers, maternity, and sub-district hospitals. The Health Centre is situated in the northeastern part of Nairobi, Kenya, and serves a low-income resident population that largely depends on public health services offered at minimal or no cost to the residents. This area comprises both lower middle-class areas and slum-type regions known as (Kariobangi North) with a population of 18,903 residents. The health center plays a vital role in the community’s well-being [27]. Similarly, Kangemi Health Center, the second study site, is a level three facility managed by Nairobi City County Government. The facility occupies a small valley on the outskirts of the city, and according to the 2019 census, Kangemi is home to 116,710 residents [27]. Therefore, this health facility serves a large informal settlement community, catering to their medical needs at minimal or no cost.

### Procedures

Community health volunteers, currently referred to as community health promoters (CHPs), recruited community members to participate in the mental health survey. The CHPs obtained consent from the participants as well as assent from minors and caregivers’ consent where it was required. The baseline data was collected through a survey administered by the CHPs. Participants identified as having moderate to severe depression were invited to attend a short therapy program. This therapy consisted of 4 sessions that were held once per week. The group sessions were facilitated by CHPs who had received training as facilitators of group Interpersonal psychotherapy. At the beginning of each session, the participants were assessed for psychological distress by the CHP facilitator. This screening was aimed to monitor their progress and help facilitators provide appropriate support throughout the therapy. Upon completion of the four sessions, the participants were screened again using the same tools used for baseline assessment. This intervention screening took place one month after the therapy had ended. The purpose of the screening was to evaluate the progress and any changes in the mental health status

### Ethical approval

Ethical approval was received from the Kenyatta National Hospital/University of Nairobi Ethical Review Committee (No. P694/09/2018) and a research permit from Kenya National Commission for Science, Technology and Innovation (NACOSTI/P/19/77705/28063). Permission was also sought from these study sites through the Nairobi County Directorate of Health (Approval no. CMO/NRB/OPR/VOL1/2019/04). This study was nested under MK’s work on Implementing mental health interventions for pregnant adolescents in primary care LMIC settings (INSPIRE) study. Written consent was obtained from participants, as well as informed assent, which was followed by seeking parental/caregiver consent for participants who were below 18 years old. Participants were assigned unique identifiers to protect their identities, collected data was stored in password-protected computers, while consent and assent forms or any document containing identifying information were kept in lock and key cabinets only accessed by authorized study staff.

### Training of facilitators

MK developed an outline of the four-session intervention by utilizing qualitative evidence that delved into the mental well-being of peripartum adolescents, with a specific emphasis on four IPT problem areas during the formative phase; the researchers were able to gain a richer understanding [13,28]. The interpersonal framework revealed crucial insights into adolescent depression, indicating its potential to guide the development of strategies to relieve their distress [14,15,29]. The implementation of this 4-session IPT-G mini was a collaborative effort. OY, a trainer in IPT-G with a background in clinical psychology and VN trained in carrying out IPT assessments and program delivery in community settings, developed standard operating procedures. Training of CHPs and intervention delivery procedures were further developed with JK and SM, who are clinical psychologists and county staff and were undergoing training to become IPT field supervisors. They carefully reviewed the overall mental and physical health needs of recruited participants. MK and OY conducted training for the CHPs on delivering the IPT-G mini. We adhered to the mini IPT protocol that was recently published, which outlined the challenges faced by peripartum adolescents in Kenyan primary care settings while also exploring strategies to strike a balance in addressing issues of adaptations, fidelity, and modification of the intervention [29]. A rapid online workshop over five days virtually provided readiness for the intervention delivery. This was followed by a workshop on refinement of intervention implementation protocol based on training feedback from CHPs.

The aim was to create an engaging and therapeutic group process where participants felt at ease and the group context accelerated social contact and the opportunity to learn. This efficacy trial provided group IPT-G mini for 40 participants experiencing significant distress, as identified through our survey.

### Intervention delivery process

The trained IPT facilitators delivered the intervention according to the IPT-G Mini protocol, receiving on-site supervision from IPT supervisors. Sessions were held weekly for four weeks, each lasting 90 minutes (Fig 2).

**Fig 2.**
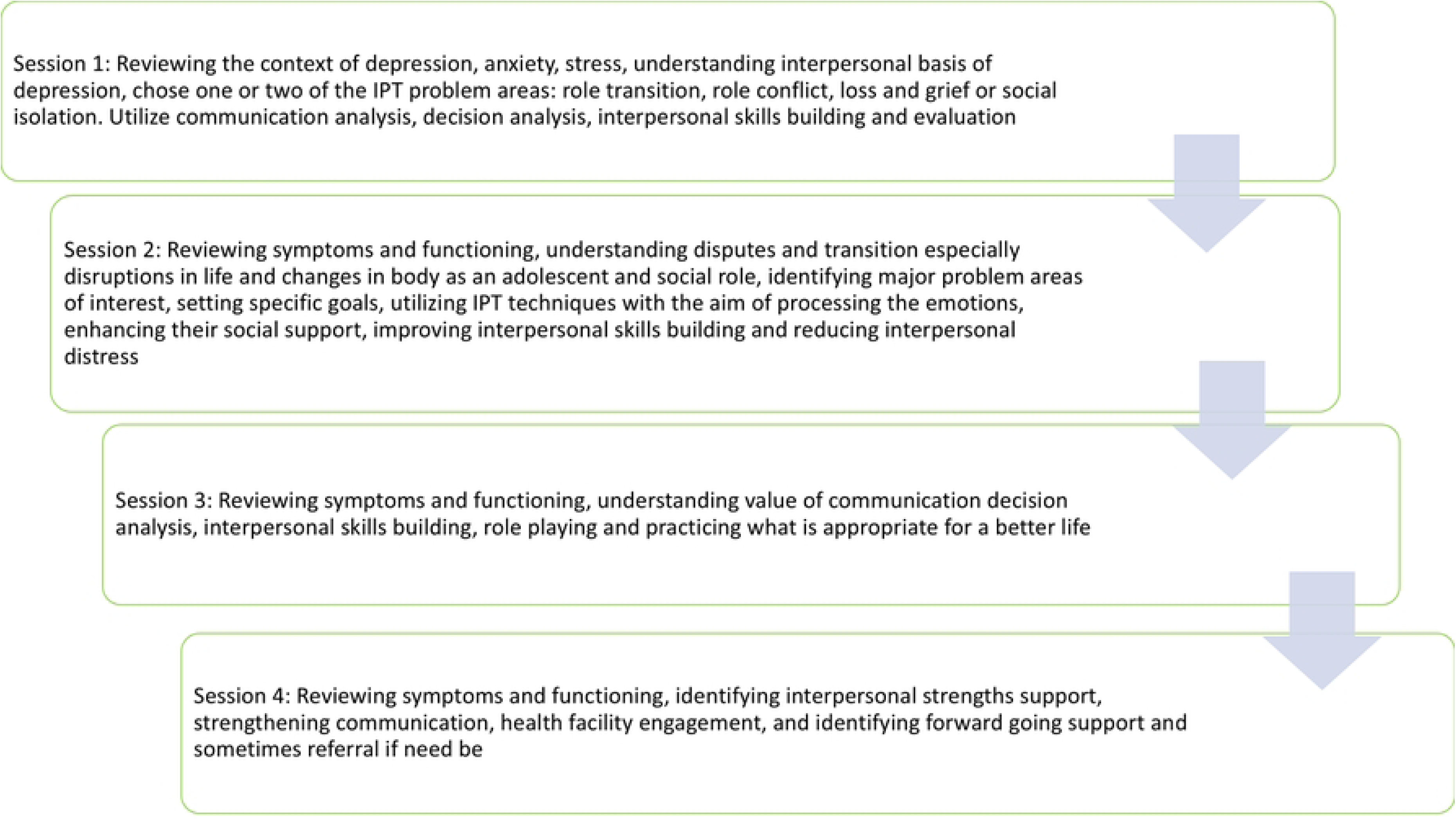
Structure of the IPT-G.

The participants were grouped based on age: 10-14 years -formed one group, while 15–19 years old formed another group. This age-based grouping aimed at providing therapeutic approaches unique to each age group’s needs and developmental stages. Importantly, grouping participants by age was a deliberate design decision and not a post-hoc arrangement. Each therapy group underwent the intervention based on their respective age categories, ensuring a focused and personalized therapeutic experience. Additionally, we offered brief psycho-educational support to the caregivers of these participants, equipping them with valuable tools to enhance the support they provide to the adolescents at home.

### Data collection

Data was collected using tablets, with baseline information on the participants retrieved from a previous MMAP survey that identified them as high risk for psychological distress. In June and July 2021, CHPs conducted baseline data collection under the supervision of team members VN and AT. The CHPs received three days of intensive training on assessment tools, ethical considerations when dealing with adolescents, and general guidelines for data protocols.

### Measures

#### Patient Health Questionnaire-9 (PHQ-9)

This is a nine-item self-report scale consisting of nine criteria used for diagnosis of depression in the Diagnostic and Statistical Manual of Mental Disorders (DSM-5) [30,31]. This scale is derived from the primary care evaluation of mental disorders, an instrument developed to assist primary care clinicians in making diagnoses of the most common mental disorders [20]. This nine-item scale is among the tools included in the MMAP protocol for assessing symptoms of depression among adolescents. The tool was developed for screening and assessing the severity of depressive symptoms in both clinical and research contexts [30].

#### Kessler psychological distress scale (K10)

This ten-item scale was developed using sensitive items that identify psychological distress in the general population and measures the frequency with which symptoms are experienced within a month [26]. These symptoms range from nervousness, sadness, tiredness, hopelessness, and worthlessness, with responses adopting a 5-point Likert scale where 1 (None of the time) and 5 (All of the time). High scores point to psychological distress, with research suggesting a cut-off point of 24 and above for psychological distress [32].

#### Revised Children’s Anxiety and Depression Scale (RCADS)

This tool follows the Spence Children’s Anxiety Scale [33] with a few more items on general anxiety and major depression symptoms per the Diagnostic and Statistical Manual of Mental Disorders, Fifth Edition (DSM-V) criteria [31]. It was initially designed as a 47-item questionnaire for ages 8–18, but a shorter 25-item version was then developed [34] which has since been validated and shortened to 22 items that demonstrated acceptable categorization for 10-14 and 15-19-year-olds [35]. We are reporting on RCADS anxiety items here as the reliability of the anxiety subscale was the most robust.

### Data analysis

Outcome measures were collected at four distinct time points: baseline, post-intervention (after 4 sessions), and 1-month post-intervention. The results are presented as means and standard deviations (means ± SD), along with effect sizes calculated using Cohen’s d. To assess group disparities, exploratory longitudinal regression analyses were conducted for each outcome. All analysis was conducted using SPSS v25.

## Results

### Socio-demographic Characteristics (N= 40)

Overall, participants were predominantly females (n=26; 65%). The mean age was 15.2 years (SD=2.9), with the majority (n=24; 60%) aged between 15-19 years. A large proportion of the respondents (n=31; 78%) resided with their parents. The majority (n=22; 55%) were in primary school, 40% were in secondary school, and 5% were engaged in vocational and livelihood programs at the tertiary level. Most respondents (40%) reported an average monthly income between Kenyan shilling (Kshs). 5,000-9,999 (50-100 USD per month). About 38% reported their average income to be Kshs. <5000 and 23% had an average income of Kshs. 10,000 and above (100 USD per month and over that). Baseline characteristics of the trial population by age were comparable between age groups (See Table 1).

**Table 1.**
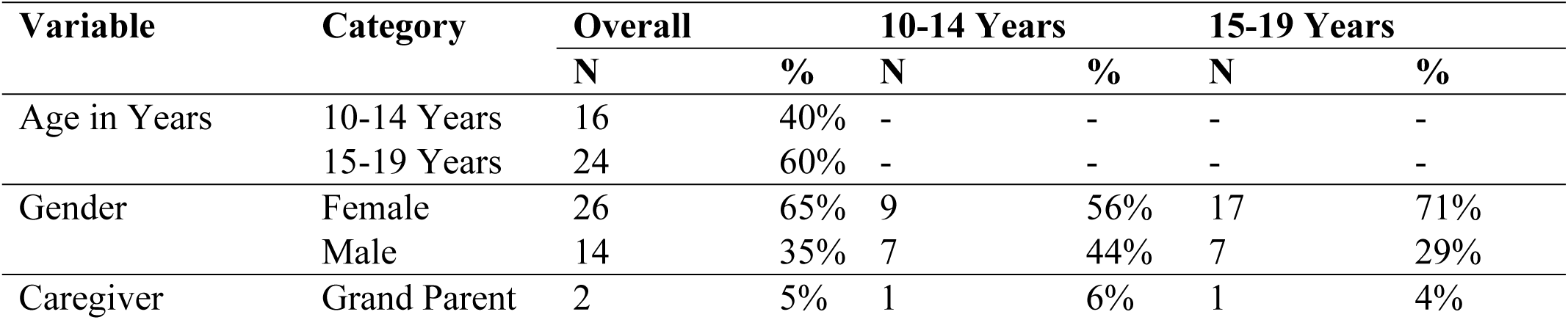
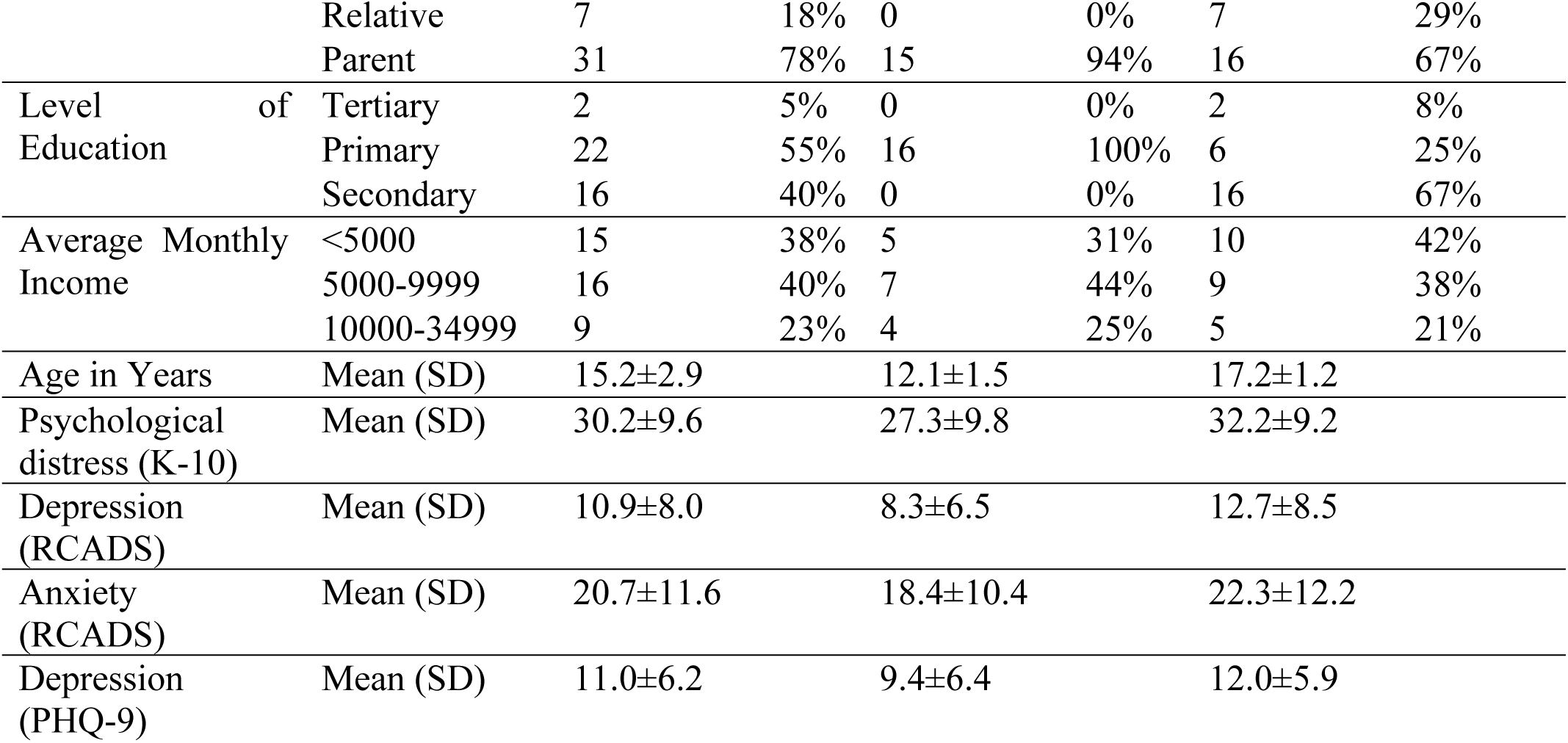
Baseline characteristics segregated by ages.

### Outcome measures

Attendance for each session was tracked by CHPs, who counted the number of participants assessed using the K10 scale, which was administered during every session. In the initial session, 40 participants attended a group session; in the second session, 37 participants attended; in the third session, 38 participants attended; while in the final (4^th^) session, 40 participants attended groups. In total, 4 groups were created, with each site having two groups. Following the intervention, there was a notable decrease in psychological distress (K-10) and depression score (PHQ-9) with a medium effect size of Cohen’ [*d=0.49*; 95% C.I. =0.04-0.94] for psychological distress and depression [*d=0.51*; 95% C.I.= 0.06-0.96] respectively. (See Table 2 for comparisons).

**Table 2.**
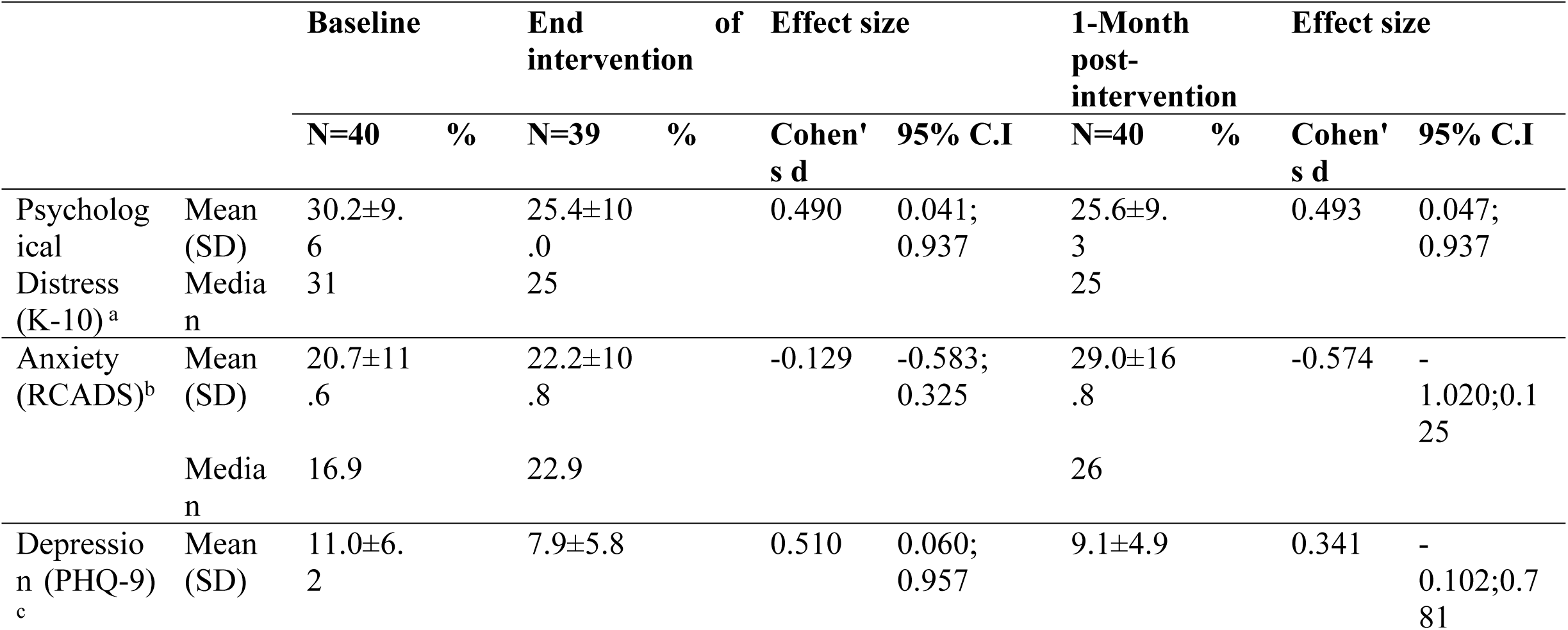
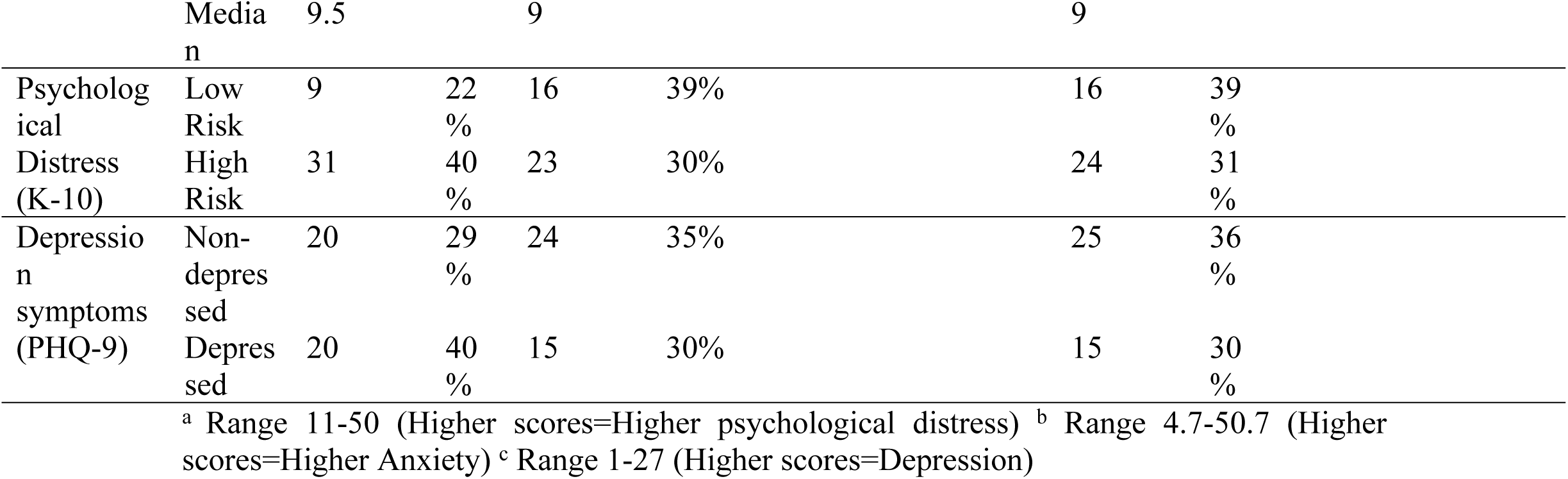
Intervention outcomes.

However, there was a non-significant increase in anxiety (on RCADS) that we noticed post-intervention. The percentage of respondents at heightened risk of psychological distress decreased from 40% at baseline to 30% at the end of the intervention and was found to be 31% at one month post-intervention. Similarly, the percentage of respondents with probable depression decreased from 40% at baseline to 30% at the end of intervention and remained the same at 1-month post-intervention.

The analysis of group variances from the exploratory longitudinal regression analyses for each outcome (See Table 3) indicated a significant decline in psychological distress (as measured by K-10) and Depression (as measured by PHQ-9) scores at the end of the intervention *p<0.05*. Notably, younger participants aged (10-14 years) exhibited more favorable outcomes compared to older adolescents on both psychological distress and depression.

**Table 3.**
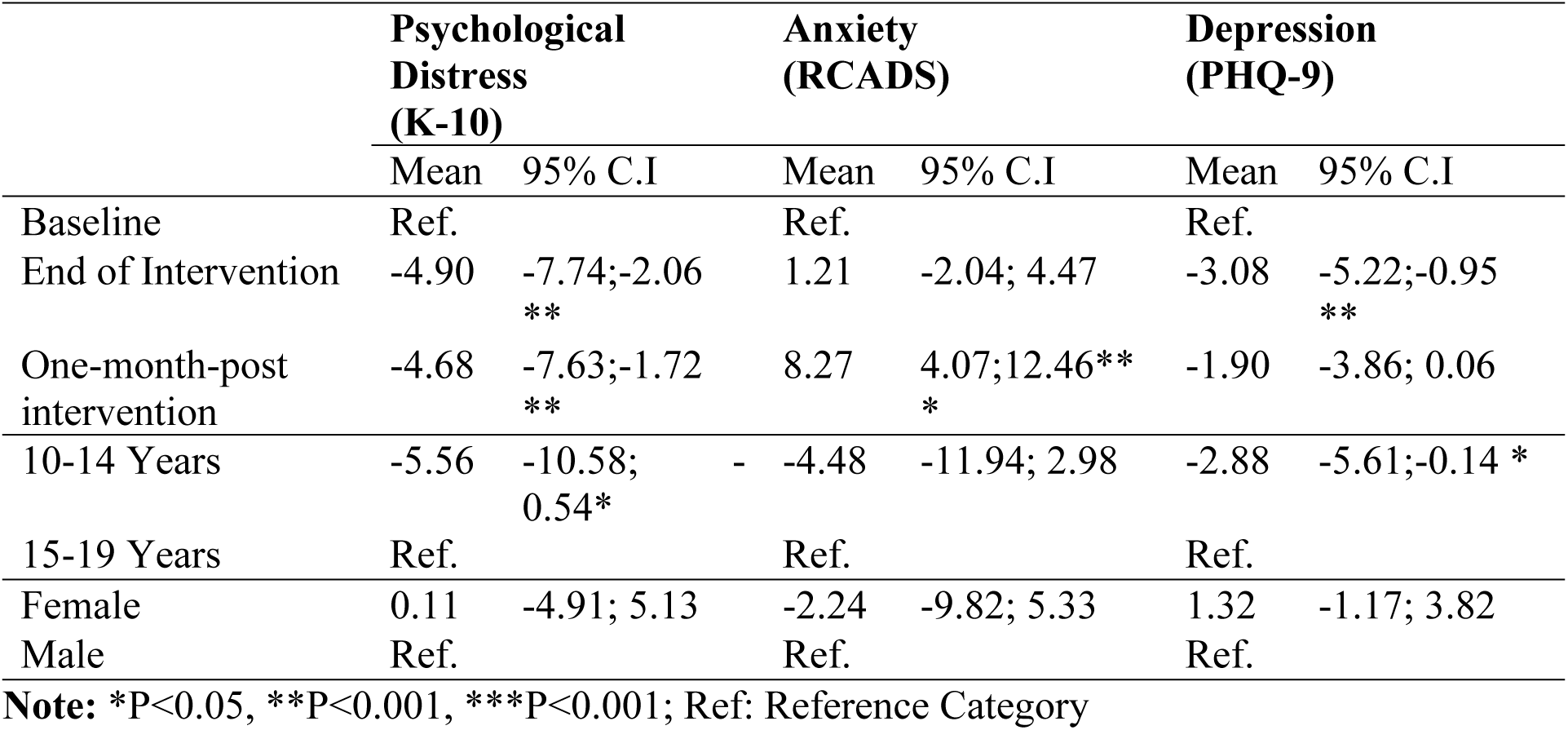
Summary of mean group differences for outcome measures.

During the one-month follow-up after the intervention, we found both clinically and statistically significant reductions in psychological distress (Cohen’s d=0.49; 95% C.I.=0.05-0.94), while depression scores showed a statistically non-significant decrease. However, the IPT team considered participant improvements to be clinically significant. We did, however, find an increase in RCADS anxiety scores.

### Effects of Intervention after each IPT session

To explore the turning point at which significant changes occurred in the outcome measures following the intervention sessions, we performed pre- and post-intervention analyses testing psychological distress from session one to one month after the intervention while controlling for age and gender (See Table 4).

**Table 4.** Effects of Intervention per session.

| Parameter | $\beta$ | S.E. | 95% C.I. | Sig. |
| --- | --- | --- | --- | --- |
| One-Month Post | -4.68 | 1.51 | -7.63; -1.72 | **0.002** |
| Termination | -4.90 | 1.45 | -7.74; -2.07 | **0.001** |
| Session 3 | -0.91 | 1.46 | -3.77; 1.95 | 0.534 |
| Session 2 | -0.99 | 1.44 | -3.82; 1.84 | 0.493 |
| Session 1 | -1.88 | 1.72 | -5.25; 1.50 | 0.277 |
| Baseline | Ref. |  |  |  |
| 15-19 Years | 5.09 | 2.38 | 0.42; 9.76 | **0.033** |
| 10-14 Years | Ref. |  |  |  |
| Male | 0.99 | 2.39 | -3.69; 5.68 | 0.677 |
| Female | Ref. |  |  |  |

$\beta$ : Regression Coefficient; S.E: Standard Error; C.I.- Confidence Interval; Ref: Reference Category

The analysis revealed a consistent decrease in psychological distress with each session, showing a marked reduction specifically at the fourth session. This decrease in distress persisted even one month after the intervention’s completion. We therefore conclude that statistically significant changes were discernible after the completion of four sessions (Figs 3 and 4).

**Fig 3.**
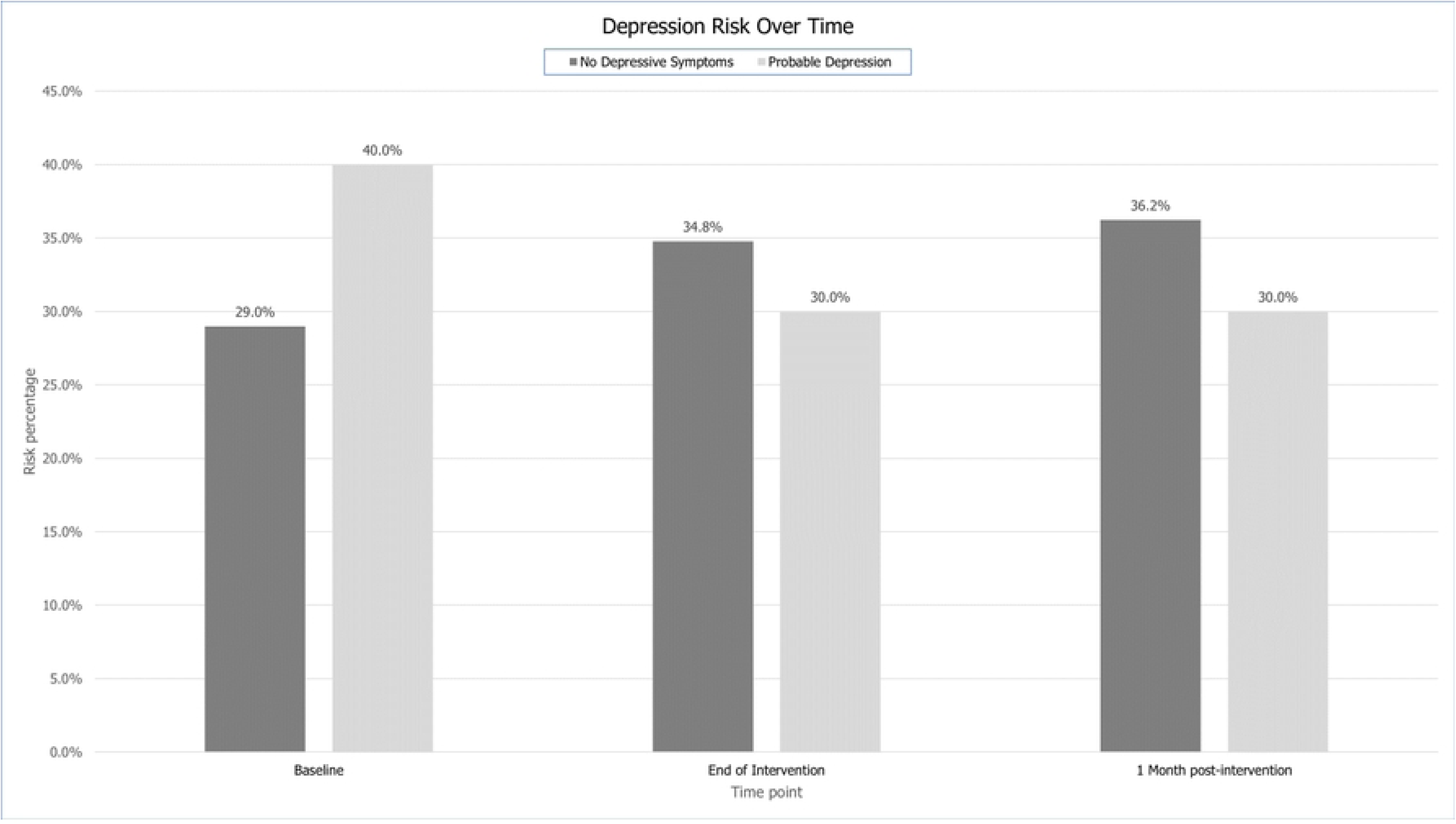
Depression severity over time on PHQ-9.

**Fig 4.**
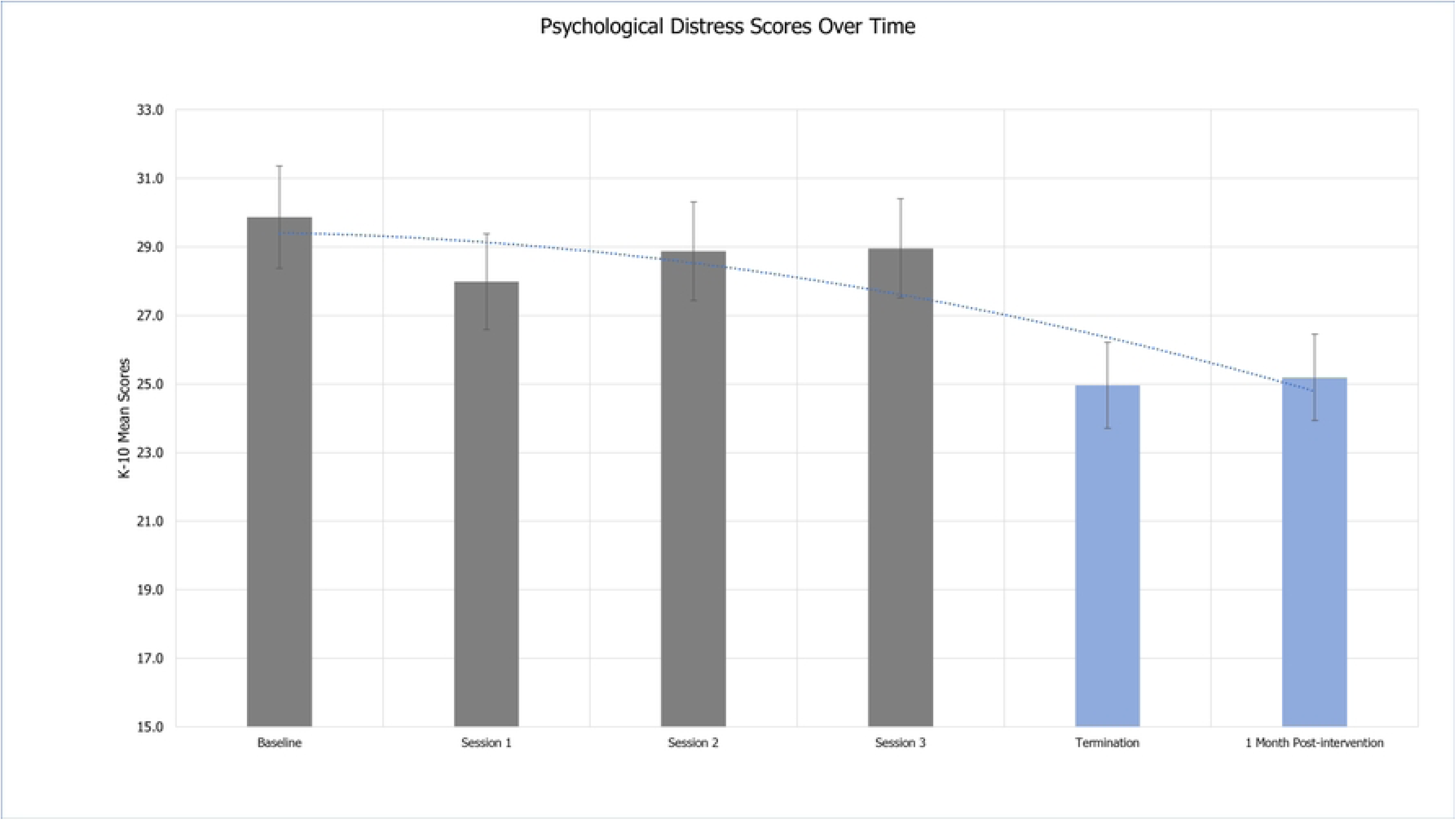
Psychological distress over time.

## Discussion

IPT-G mini effectively reduced the levels of psychological distress and depression among our young participants. These positive effects were sustained for up to one month post-intervention. However, a noteworthy increase in anxiety scores following the sessions prompts further investigation. We would also like to underscore that the intervention was delivered at the pandemic’s peak. As a result, communities were experiencing high levels of distress and uncertainty. There are a few explanations underpinning our findings. We suspect that mixing participants of somewhat different ages and varying severities of symptoms in groups may have led to the emergence of anxiety symptoms, particularly among those recovering from severe comorbid symptoms associated with grief and trauma. We also noted that at the start of the sessions, there was a rise in anxiety levels, and this could be attributed to various factors like the ongoing pandemic amplified fears and worries among the adolescents, lack of basic needs like food, insecurity, escalating poverty, and parental well-being, which were not all adequately addressed in our intervention as this was a complex, evolving situation everywhere with lockdown and national economic stress. Additionally, after completing the four sessions, we also observed triggers for anxiety was the termination of the session as participants expressed their desire to extend their time together. Towards the end of the session participants exhibited separation anxiety as they had begun to form attachment with the group and facilitators. Fortunately, CHPs could follow up with participants, and the study team also provided information to caregivers for additional support during the pandemic. No adverse events were encountered during this trial.

Our participants were experiencing a range of challenges. The ongoing stress of the COVID-19 pandemic, coupled with unprocessed grief arising from abusive family dynamics, contributed to significant distress. Insights from prior focus group discussions with adolescents aged 10-14 and 15-19 years, as well as caregivers of 10-14 years old, [24,36], revealed poignant stories. Adolescent girls disclosed accounts of contemplating suicide in relation to their experience of unplanned pregnancy, sometimes due to coercive sex, and shared attempting unsuccessful abortion. These problems around adolescent sexual and reproductive health, rights, and safety are well-documented, and during the pandemic, the closure of schools for long periods contributed to significant distress for families, children, and adolescents with limited resources [36].

### Implications of our IPT-G-Mini trial

Earlier studies [13,37–39] have indicated that briefer therapeutic interventions are well-received and viable [11,40]. Adini et al., determined that Interpersonal Psychotherapy-Adolescent-suicide crisis intervention (IPT-A-SCI) is feasible and as effective as the standard therapy in alleviating suicidal tendencies, depression, and anxiety symptoms in children and adolescents [28].

The utilization of lay health workers known as CHPs in the Kenyan context-some of whom are affiliated with health care facilities while others are embedded within community settings-presents a valuable transition from using the workforce from formal to informal care, bringing psychosocial interventions to communities and adolescents during times of need. Our study benefitted from the seamless network of these health workers, as evidenced by the lack of dropouts among our participants.

In light of the pressing need for prompt interventions, we organized our study participants into groups based on ease of access to the facility from their homes. Group therapy sessions were conducted at their nearest primary care center. The therapy was well-received and provided relief by alleviating symptoms of depression, speaking to suicidal ideations, grief, and loss feelings, and relieved pandemic-associated worries, isolation, and social disconnect through enhancing and leveraging group support in a safe environment.

## Limitations

The primary constraint of our study included limited sample size and the absence of a control group or comparison group. This was happening during the real-world constraints of the pandemic. Provision of timely support to selected adolescents in high need was an advantage despite these limitations, our implementation of IPT -G Mini proved to be safe, feasible and highly acceptable to participants and our lay CHPs who delivered the intervention.

## Conclusions

The study demonstrates the effectiveness of IPT-G mini sessions in significantly reducing psychological distress among our young participants, with these positive effects lasting for up to a month after intervention. Nonetheless, the noted increase in anxiety scores warrants further examination. This increase may be linked to individuals with higher anxiety symptoms and may be a result of participants recovering from unaddressed severe comorbid psychopathologies such as trauma exposures, which were not specifically addressed during the intervention. It is important to acknowledge that the intervention coincided with the peak of the COVID-19 pandemic, a time when communities were experiencing high levels of distress and uncertainties. Our findings suggest IPT-G mini needs to be tested further in controlled conditions to examine its effectiveness further for a general population of adults and adolescents.

## Data Availability

The datasets generated and/or analyzed during the current study are publicly available from the corresponding author upon request

## Acknowledgments

The authors would like to thank all the participants, our facilitators, Eunice Oalo, Edith Apondi, Ann Nyaroche, Josphat Asande, Phyllis Njoki, the Nairobi County health directorate, the Director of Mental Health, the Ministry of Health, Kariobangi, and the Kangemi health facility staff.

